# Wilson’s Central Terminal Changes Location on the Body Surface During the P-Wave: Why Precordial Leads Might Not Be What We Think

**DOI:** 10.64898/2026.05.20.26352966

**Authors:** Jule Bender, Job Stoks, Cristian Barrios Espinosa, Silvia Becker, Matthijs J.M. Cluitmans, Axel Loewe

## Abstract

**Background and Aims:** Clinical interpretation of the precordial leads V1-V6 assumes that Wilson’s central terminal (WCT) has a fixed anatomical location. Consequently, a positive signal corresponds to electrical activation spreading from WCT towards the respective electrode, and vice versa. However, the location of WCT has never been systematically investigated. Yet, a better understanding of WCT location could improve the interpretation of the precordial leads. This work aims to characterize the spatial expansion and location of the physical WCT i.e., the electrical potential defined by the WCT, during the P-wave on the body surface.

**Methods:** An intensive analysis of body surface potential maps (BSPMs) during atrial depolarization in an in silico patient cohort and clinical data was conducted.

**Results:** During the P-wave, the location of WCT was not stationary but the spatial extent and location varied across time as well as across individuals. Four distinct spatial patterns of WCT distribution on the body surface were identified in silico, and three of these were found in the clinical cohort. WCT signals agreed with BSPM signals at commonly assumed positions of WCT only for a small fraction of the P-wave.

**Conclusion:** The spatial extension and location of WCT changes during the P-wave and thus should be considered when interpreting the precordial leads.

**Translational perspective:** This study highlighted that the spatial extent and location of Wilson’s central terminal (WCT) on the body surface change during the P-wave. Variability in time as well as across individuals suggest that assumptions on a fixed WCT location, such as for the precordial leads, should be treated with caution.

## 1 Introduction

The 12-lead electrocardiogram (ECG) is a widely used diagnostic tool in cardiology due to non-invasiveness, standardized acquisition, routine availability, and low cost. Of the 12 leads, the unipolar leads V1-V6 utilize Wilson’s central terminal (WCT) as a virtual reference electrode. Thereby, it is assumed that WCT is a passive reference with a potential close to zero [1, 2]. Specifically, the WCT potential is obtained by averaging the electrical potentials of the right arm *ϕ*_RA_, left arm *ϕ*_LA_, and the left leg *ϕ*_LL_ extremity electrodes to

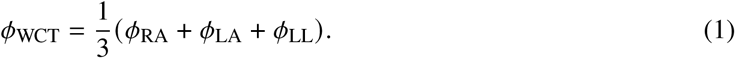

The leads V1-V6 are then defined as the potential difference between each of the six electrodes placed on the chest and the potential *ϕ*_WCT_ [1]. Conceptually, these leads are interpreted as vectors pointing from WCT inside the chest towards the corresponding chest electrodes, providing a horizontal view of the electrical depolarization of the heart. Thus, a positive deflection in the precordial leads reflects electrical activation spreading from WCT towards the corresponding electrode, and vice versa. Understanding WCT is crucial for interpreting the leads V1-V6 in both clinical practice and theoretical analysis.

Although several experimental and numerical studies [3, 4, 5, 6, 7, 8, 9] have challenged the concept of WCT by showing that it is neither zero nor steady during the cardiac cycle, it was standardized as reference electrode for the precordial leads in 1954 [10]. Recent measurements of “true unipolar leads” [11] confirmed that WCT amplitude is non-negligible, time-dependent, may exceed P-wave amplitude, varies between individuals, and can dominate Wilson lead morphology, potentially decreasing sensitivity and specificity [3]. Despite these findings, the clinical utility of the precordial leads have led to a largely unchallenged, widely accepted use and limited research on WCT [3]. Consequently, when interpreting the Wilson leads, WCT is still considered to have zero potential and a fixed location [12]. The location of this anatomical WCT is typically assumed as the center of the chest, between V1 and V2 electrodes or at the center of Einthoven’s triangle [2, 12, 13, 14]. However, the true location of the physical WCT, i.e. the location of the electrical potential defined by the WCT, has never been systematically investigated [3] and recent work suggests that the Einthoven triangle cannot always be identified as limb-lead voltages form a triangle only during parts of the QRS complex [15]. In addition, in practice, the interpretation of the Wilson leads can be challenging and some ECG markers based on these perform worse than expected. This challenge is exemplified by P wave terminal force in lead V1 (PTF-V1), a marker characterizing the terminal portion of the P-wave in lead V1, used to diagnose left atrial enlargement (LAE) [16]. Theoretically, a late negative P-wave with high amplitude and a large PTF-V1 is expected for LAE, as left atrial activation points away from the center of the chest. However, in practice, relatively low sensitivity and specificity are observed [16, 17, 18, 19, 20, 21, 22] which may be partially attributed to misinterpretation of the reference electrode.

This work aims to investigate the location of the physical WCT, i.e. the electrical potential defined by the WCT, on the body surface during atrial depolarization. We hypothesize that the spatial expansion and location of the physical WCT vary during the cardiac cycle. To this end, surface regions where the WCT potential matches the body surface potential map (BSPM) were identified with a large in silico study considering the entire torso surface, followed by confirmation in clinical data.

## 2 Methods

This study combines two approaches: an intensive analysis of simulated BSPMs during atrial depolarization in an in silico patient cohort and clinical data in a smaller cohort. The methodology used is visualized in Figure 2 and explained in detail in the Supplementary Material A.

**Figure 1.**
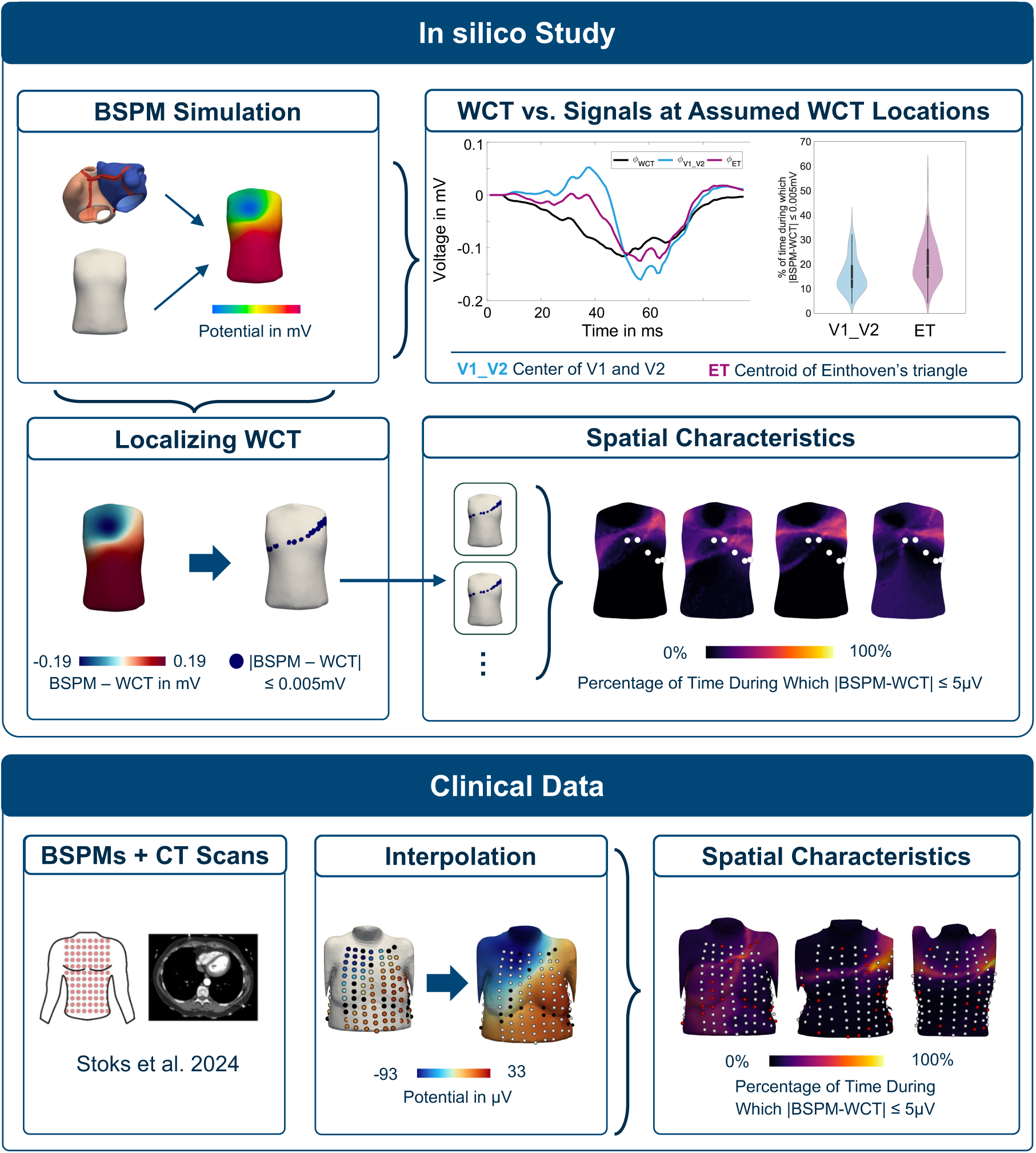
Graphical Abstract.

**Figure 2.**
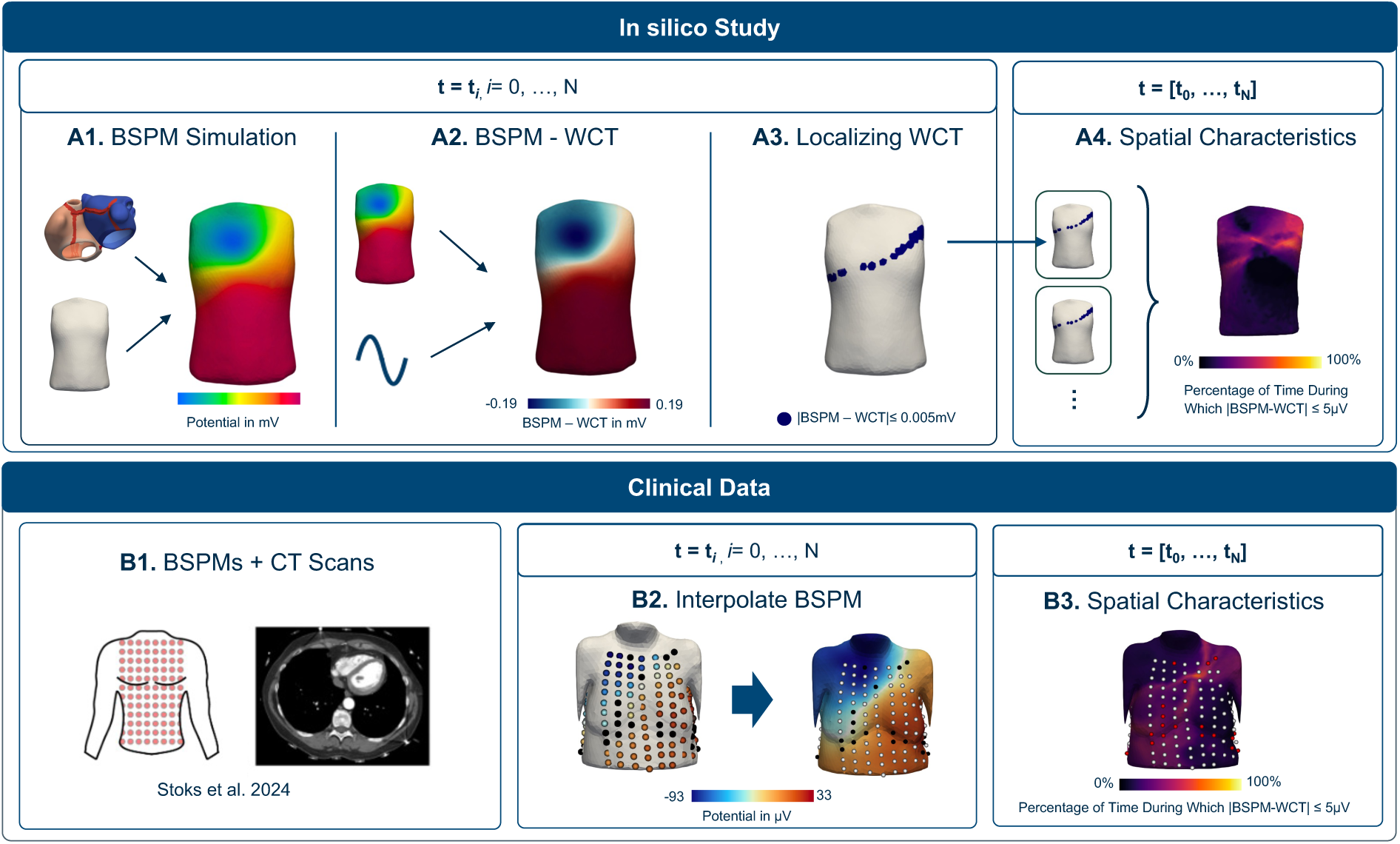
Study methodology: **Top:** In silico study to analyze the location of WCT. **A1:** Simulation of BSPMs for each timestep using an atrial and a torso model. **A2:** Computation of WCT and the difference between the BSPM and WCT for each time step *t*_*i*_. **A3:** Localizing WCT on the body surface for each time step *t*_*i*_ using a fixed threshold. **A4:** Analysis of spatial histograms of WCT during the entire P-wave ([*t*_0_,…, *t*_N_] and manual pattern classification. **Bottom:** Analysis of WCT location in the clinical data. **B1:** Body surface potential measurement and CT scan of the torso were acquired in a previous study [23]. **B2:** Interpolation of the BSPM to the torso models derived from the CT scan. **B3:** Analysis of WCT location as in panels A2, A3, and A4. WCT: Wilson’s central terminal, BSPM: Body surface potential map.

### In silico Study on WCT Location

A virtual cohort was created based on 100 bi-atrial and 25 torso geometries using statistical shape models (SSMs) [24, 25, 26]. Body surface potential maps during atrial depolarization in sinus rhythm were simulated using the Eikonal equation and the boundary element method [24, 27]. The overall spatial distribution of WCT during the P-wave was characterized by the percentage of time *p* during which the absolute difference of WCT and BSPM was smaller than 5 *μ*V (see Supplementary Figure S1). The resulting spatial distributions were manually classified. Additionally, WCT was compared to the BSPM signals at the center of the Einthoven triangle projected on the body surface and the center between the V1 and V2 electrode positions.

### Clinical Data on WCT Location

For clinical analysis, BSPMs from 10 normal subjects (30 % male) acquired previously [23] were analyzed (Table 1). All patients had structurally normal heart on echocardiogram, normal 12-lead ECG, and no (suspected) pathology affecting ventricular electrophysiology. This study was approved by the local ethics committees of Maastricht University Medical Center (MUMC), The Netherlands (METC 11-2-043), and adhered to the Declaration of Helsinki. All subjects provided written informed consent prior to inclusion. Each BSPM was recorded using approximately 185 electrodes at a sampling frequency of 2048 Hz. For the analysis, 10 s segments of the BSPMs were extracted, noisy signals were semi-automatically identified and removed [23], and P-wave templates were generated [28]. P-wave onsets and offsets were then annotated manually based on the 12-lead ECG template [28] and all BSPM templates were bandpass filtered (0.05-40 Hz) and corrected for isoline offset. Additionally, non-contrast low-dose thoracic CT scans were available to identify the electrode positions [23]. As in silico study, the percentage of time *p* during which BSPM and WCT coincide up to a tolerance was determined after interpolation to the CT scan of the torso surface (see Supplementary Figure S1). For each of the 10 individuals, the spatial pattern of WCT was identified manually based on the categories defined based on the results of the in silico study.

**Table 1.** Patient characteristics of the clinical cohort [23]. F: female, M: male.

| Sex | Height (cm) | Weight (kg) |
| --- | --- | --- |
| F | 169 | 75 |
| F | 163 | 65 |
| M | 170 | 70 |
| F | 177 | 128 |
| F | 170 | 60 |
| F | 176 | 79 |
| F | 172 | 91 |
| F | 165 | 54 |
| M | 186 | 90 |
| M | 176 | 62 |

## 3 Results

### WCT Signal Differs from Those at the Assumed Location

WCT morphology and amplitude differed from the BSPM signals at both the center of the Einthoven triangle, *ϕ*_ET_(*t*), and the center between the electrodes V1 and V2 *ϕ*_V1___V2_(*t*) (Figure 3 A). Overall, WCT was not constant during the P-wave but exhibited time-dependent amplitude changes that followed the temporal course of the P-wave (Figure 3 A). In total, the WCT signal agreed with BSPM signals at commonly assumed anatomical positions of WCT only for a small fraction of the P-wave. Across all atrial geometries combined with the mean torso model, the average agreement was 15.8 % and 20.9 % for the center of the electrodes V1 and V2 and for the centroid of Einthoven’s triangle, respectively. Few models showed higher agreement up to 57 % during the P-wave (Figure 3 B). For different torso models and the mean atrial model, mean agreement was 11.7 % and 16.4 % for the center of V1 and V2 electrodes and for the center of Einthoven’s triangle, respectively. However, for some models, agreement up to 35 % could be observed for these points (Figure 3 C).

**Figure 3.**
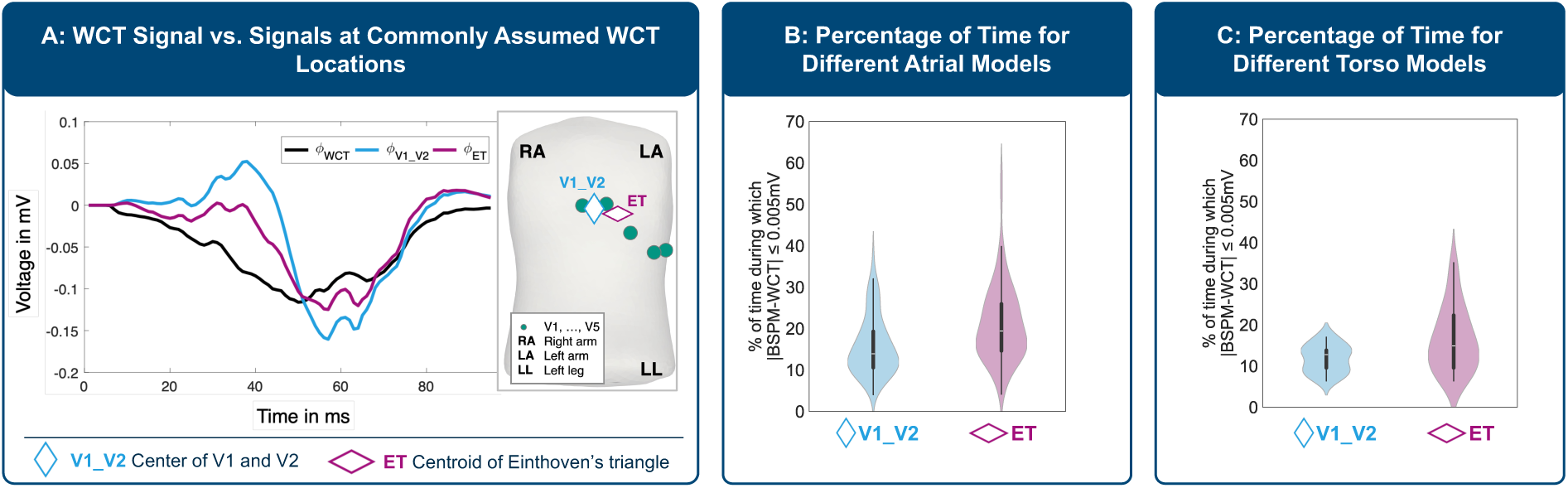
Comparison of WCT signal and signals at commonly assumed WCT locations. **A:** Signals of mean atrial and torso geometries. WCT is visualized in black, the signal *ϕ*_V1 V2_ at the center between the V1 and V2 electrode positions in blue, and the signal *ϕ*_ET_ at the center of Einthoven’s triangle on the body surface in purple. **B:**Distribution of time during which the signals coincided for 99 atrial models combined with the mean torso model. **C:** Distribution of time for mean atrial model combined with 25 torso models. WCT: Wilson’s central terminal, RA: right arm, LA: left arm, LL: left leg, ET: Einthoven’s triangle.

### WCT Changes Location During P-Wave

During the P-wave, the physical WCT location was not stationary. Consequently, no point was found at which WCT was located during the entire P-wave, with a maximum of 73 % agreement observed for individual geometries in silico. Although the spatial distribution varied over time in all models, in general WCT was located superior to the precordial electrodes of the standard 12-lead setup, typically on the left lateral chest. For the mean atrial and torso geometries, WCT first extended from the right shoulder to the left chest, then became more horizontally oriented, and extended from the left shoulder to the right chest during the terminal phase of the P-wave (Figure 4, white area and Supplementary Figure S4 and Supplementary Videos 1-5.).

**Figure 4.**
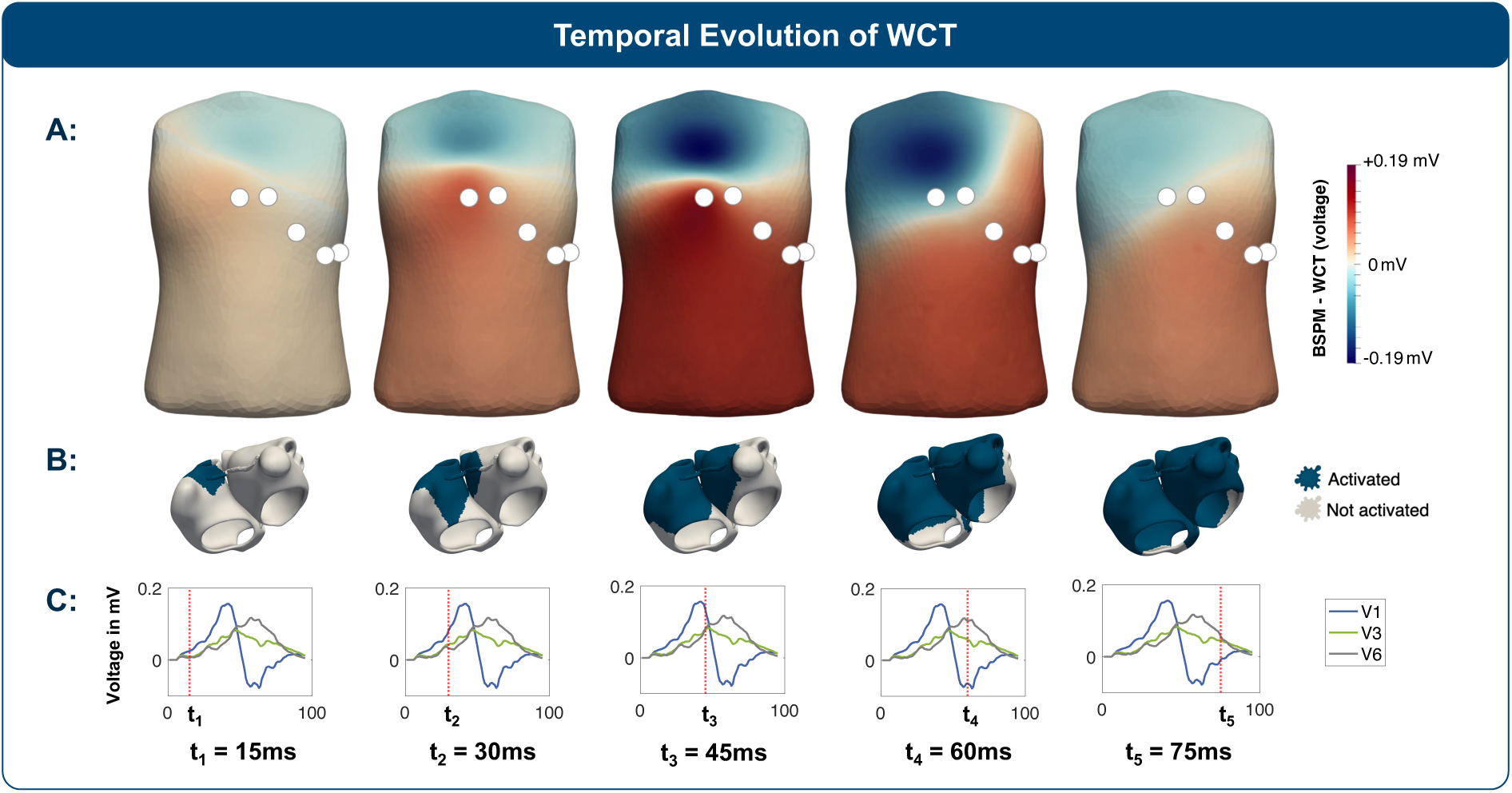
**A:** Temporal evolution of the WCT location on the body surface for the mean atria and torso model. Electrode positions for leads V1-V5 are indicated by white spheres. White areas on the body surface indicate regions where the absolute difference between WCT and the BSPM was ≤ 5 *μV*, and can thus be considered as WCT location. **B:** Depolarized regions of the atria at each time point are shown in blue. **C:** Signals of the leads V1, V3, and V6 with red vertical lines marking the corresponding time step. WCT: Wilson’s central terminal, BSPM: Body surface potential map.

### Different Spatial Patterns for WCT

Four distinct spatial patterns of physical WCT distribution on the body surface were identified in silico, three out of these four were also observed in the clinical cohort (Figure 5 and Supplementary Figure S4). In silico, for different atrial geometries combined with the mean torso model, most of the models could be assigned to pattern A, characterized by WCT extending from the left shoulder towards the right chest (33 models), and pattern B, extending pattern A from the right shoulder toward the left chest (28 models). Moreover, 16 models were assigned to pattern C, characterized by high percentages on the left chest only and 19 models were assigned to pattern D, showing a horizontal distribution of WCT location from the left to the right chest. Three models had large percentages in the abdominal region and could not be assigned to one of the four patterns (Figure 5 and Supplemental Figure S3). Conversely, for varying torso models and the same atrial geometry, similar spatial patterns of WCT were observed (Supplementary Figure S2 A) and WCT signals exhibited similar morphology but differed in amplitude. In the clinical cohort, three patients were assigned to pattern A and C each and 4 patients to pattern D. Overall, while some models had an extended area considered as WCT location during parts of the P-wave, only a small area was detected for others. In general, differences occurred mainly on the right chest were less evident at the extremities, yielding similar WCT morphology for the four spatial patterns. Overall, no position was identified where WCT was located for more than two-thirds of the P-wave in silico. Larger chest diameters were associated with higher mean percentages for WCT locations (Supplementary Figure S2 B and S2 D). Although the union of possible WCT locations for different models covered a broad region of the thorax (Supplementary Figure S2 C), across all models, no common point was found where WCT was located for the majority of the P-wave duration. Interestingly, visually similar Wilson lead morphology was observed among the different spatial patterns.

**Figure 5.**
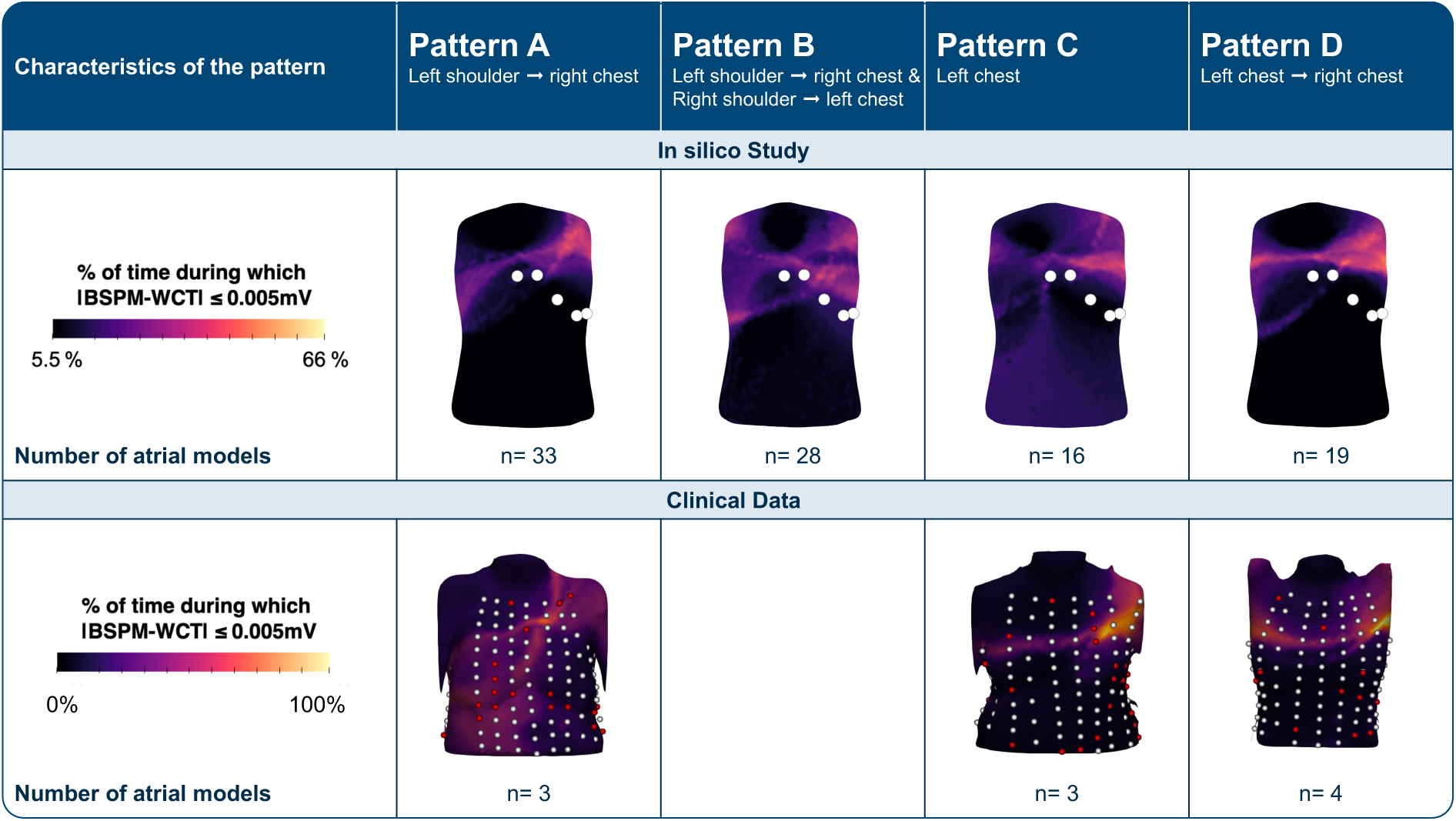
Spatial patterns of WCT on the body surface, manually classified into four different categories by visually analyzing the spatial extension and location of areas with high percentages on the body surface. 3 out of 99 models could not be assigned to one of the four patterns but had large percentages in the abdominal region. In the clinical cohort, no example for pattern B was found. Spheres represent electrode positions of the BSPM, and electrodes that were too noisy and thus not used in the analysis are visualized in red. The temporal evolution of the WCT location on the body surface for each pattern is visualized in Supplementary Figure S4. WCT: Wilson’s central terminal, BSPM: Body surface potential map.

## 4 Discussion

### WCT Location Varies over Time and Across Individuals: Implications for Interpreting Precordial Leads

An ideal unipolar reference electrode has zero amplitude, is constant, and most importantly has a fixed location [4, 14]. While time-dependent P-wave characteristics of WCT amplitude have been reported previously [3], the present study demonstrated that the spatial extent and the location of the physical WCT on the body surface vary over time and across individuals, both in silico and clinically. The results further indicate that no other unique universal position could serve as an alternative to conventionally assumed anatomical WCT locations [12]. In particular, no point on the torso surface was identified where WCT was located for at least half of the P-wave in all models, and the overall spatial extension where WCT occurred was large. Specifically, the physical WCT location did not match commonly assumed locations of anatomical WCT at the center of the body [12]. Consequently, interpreting the Wilson leads under the assumption of a fixed reference location is likely inaccurate. Therefore, any assumptions on a fixed location should be treated with caution when interpreting WCT and the precordial leads V1-V6. In this context, WCT should rather be considered as a truly virtual (i.e. not linked to a specific anatomical location), time-dependent reference electrode with non-zero potential, consistent with previous discussions [4]. Particularly, low sensitivity and specificity values for precordial lead interpretation as for PTF-V1 [16, 17, 18, 19, 20, 21, 22], may be attributed to the spatial characteristics of WCT. Due to variable WCT location, left atrial activation does not necessarily point away from the assumed anatomical WCT reference. Consequently, the magnitude of PTF-V1 may not consistently increase for LAE as currently assumed and thus may decrease the sensitivity of PTF-V1 to detect LAE. Furthermore, other components of ECG interpretation that rely on WCT, such as the right chest leads [5] or posterior leads [29], may likewise be affected by inaccurate WCT assumptions and should be interpreted with equal caution.

### Anatomy-Dependent Characteristics of WCT

While larger variability in the spatial location and extent of WCT was observed across different atrial geometries compared to variations in torso models in the simulations, visually similar Wilson lead morphology was observed among the different spatial patterns. While similar WCT signals might be attributed to less pronounced differences in the BSPM signals at the extremities, the similarity in the Wilson leads may be explained by the fact that the signals can be dominated by WCT as shown in previous work [3]. Furthermore, visual inspection showed that WCT signals were highly similar for all torso models when combined with the same atrial geometry, while the spatial patterns for different torso models varied depending on the distance between the torso and the atrial surface. Larger torso volumes yielded smaller BSPMs and WCT signals, increasing the likelihood of fulfilling the threshold criteria (Supplementary Figure S2). This aligns with previous studies showing that torso variability mainly affects P-wave amplitude [26, 30]. Consequently, our results suggest that the inter-individual variability for WCT reported in other studies [3] might be attributed to differences in atrial anatomy. To account for variations in amplitude, an amplitude-dependent threshold was alternatively evaluated for the clinical cohort. While this approach yielded lower percentages compared to a fixed threshold, the overall spatial patterns were similar but more smoothed (Supplementary Methods A.2 and Supplementary Figure S1).

### Contributions to the Debate on WCT

While the concept and interpretation of WCT have been questioned before [3], only few previous studies discussed location-related aspects for WCT [3, 7, 15, 31]. Our findings extend earlier discussions by Burger [4] and Dower et al. [5], who emphasized that WCT was never intended to be a physiologically meaningful reference but rather a convenient average of limb potentials. Accordingly, they argued that unipolar leads should be considered simply as the difference between a measurement electrode and a reference with fixed location. Such a reference electrode may be WCT, that is simply the average of three limb electrodes and is not zero [4]. Importantly, our findings dismiss the idea of using WCT as a reference electrode with fixed location [12] or a fixed direction of the WCT lead vector [7] by demonstrating that the spatial location of WCT is dynamic and patient-specific. This discrepancy likely arises as earlier studies did not account for the temporal changes in WCT observed in our analysis. Instead, our findings support the view that WCT is just a convenient reference derived from the three limb potentials. Likewise, our results align with a previous study based on ventricular activity that concluded that a center of Einthoven’s triangle cannot be identified using a similar metric as ours [15].

### Study Limitations

Knowing that the analysis of WCT is inherently a three dimensional problem [12], this study was restricted to the body surface to reduce complexity, facilitate the analysis, limit computational effort, and ensure a link to signals that can be measured in practice. Hence, while the physical WCT likely extends inside the thorax, potentially aligning with anatomical WCT locations, the surface analysis suggests that WCT is not a fixed point but rather highly variable, regardless of internal torso dynamics. Despite focusing only on anatomical variability, substantial variability was already observed in WCT location, suggesting even greater diversity when incorporating additional sex-specific, electrophysiological or pathological variability [7]. Comparison of clinical and simulated patterns was constrained by the small sample size, limited spatial resolution, signal noise, and shorter P-wave durations and thus higher percentages in the clinical data, nevertheless three of the four in silico WCT patterns could be robustly identified. Finally, this work was limited to the atria and the P-wave. Extending this work to the ventricles to assess how WCT variability affects QRST interpretation could be approached with similar methods in a future study. Similarly, the Goldberger leads, which also rely on virtual reference electrodes, could be subject of future studies.

## Conclusion

This study confirmed the hypothesis that the spatial extent and location of WCT on the body surface change during the P-wave. As variability of WCT location was observed in time as well as across individuals, the findings challenge the conventional assumption of a fixed universal anatomical WCT location and suggest that any assumptions in this regard should be interpreted with caution. Instead, variability should be considered when interpreting the precordial leads, both for theoretical reasoning and clinical diagnosis. Consequently, unipolar lead interpretation may be less straightforward than typically assumed. Furthermore, since the leads V1-V6 are well-established and have demonstrated clinical utility but their sensitivity for specific diagnoses remains limited, the spatial variability of WCT demonstrated in this study offers an explanation for these limitations. Finally, this study suggests that there may be better, yet more complex, alternatives for anatomical WCT location assumption when interpreting the precordial leads. Therefore, advanced ECG analysis methods considering a dynamic reference location to be developed in future work could potentially improve the diagnostic interpretation of the precordial leads.

## Data Availability

The simulated data underlying this article are openly available under the creative commons license CC-BY 4.0 in Zenodo, at https://dx.doi.org/10.5281/zenodo.20086105. The bi-atrial SSM and the derived models are openly available under creative commons license CC-BY 4.0 [32]. The body SSM is openly available under the Simplified BSD License at http://humanshape.mpi-inf.mpg.de/. The clinical data underlying this article are available at https://doi.org/10.1093/europace/euae172. Additional data underlying that article will be shared on reasonable request to the corresponding author, subject to applicable ethical and data protection regulations.

## CRediT Authorship Contribution Statement

J.B.: Conceptualization, Data curation, Software, Formal analysis, Investigation, Methodology Visual-ization, Writing – original draft; J.S.: Data curation, Writing – review & editing; C.B.E.: Methodology, Writing – review & editing; S.B.: Software, Writing – review & editing; M.J.M.C.: Supervision, Funding acquisition, Writing – review & editing; A.L.: Conceptualization, Methodology, Supervision, Funding acquisition, Writing – review & editing, Project administration, Funding acquisition, Resources.

## A Supplementary Methods

### A.1 In silico Study on WCT Location

#### Virtual Cohort Generation

A virtual cohort was created based on 100 atrial geometries derived from a bi-atrial SSM [1]. All geometries are publicly available [2] and include a volumetric wall with homogeneous thickness, interatrial bridges, rule-based fiber orientation, and tags for anatomical structures [1, 3]. To obtain a representative sample of the general population, the coefficients of the 24 eigenvectors in the SSM were randomly drawn from a Gaussian distribution within [−3*σ*, +3*σ*] [1]. To compute BSPMs, the atrial models were placed in 25 torso geometries derived from a human body SSM [4, 5]. For the torso models, the two leading eigenvectors of the SSM were varied systematically in a range of [−2*σ*, +2*σ*], covering 80 % of the total shape variance. The first eigenmode represents variation in abdominal size, whereas the second captures changes in the chest region.

#### Electrophysiological Simulations

Body surface potential maps during atrial depolarization in sinus rhythm were simulated using the Eikonal equation and the boundary element method (BEM) [1, 6]. Conduction velocities and anisotropy ratios for seven distinct regions in the atria were chosen as in previous studies (see Supplementary Table 2) [1]. Activation was initiated at the sino-atrial node at the junction of crista terminalis and the superior vena cava [7]. Local activation times (LATs) in the atria were computed by solving the Eikonal equation [6, 8, 9]. The spatio-temporal distribution of the transmembrane voltage in the atria was computed by shifting a pre-computed Courtemanche et al. [10] action potential template in time according to the LATs. In a subsequent step, BSPMs were computed by means of the BEM, using average mesh resolutions of 3 mm (atria) and 15 mm (torso) and conductivities of 0.1 and 0.2 S/m, respectively [11]. Laplacian smoothing was applied to ensure continuous propagation [6, 12]. The zero reference was chosen at the right leg (RL). A standard setup for the 12-lead ECG with extremity electrodes placed on the torso was used to compute WCT.

#### Localizing WCT in silico

WCT was analyzed during the P-wave, defined as the time interval between the earliest (*t*_min_) and latest (*t*_max_) LAT in the atria. WCT was computed using standard electrode positions for right arm (RA), left arm (LA) and left leg (LL) on the torso. For each torso surface point *x* and each time *t*_*i*_, the difference between the BSPM value *ϕ*(*x*, *t*_*i*_) and the WCT potential *ϕ*_*WCT*_ (*t*_*i*_) was computed. Points for which the absolute difference was below a fixed, empirically chosen threshold (*τ* = 5 *μ*V, see Supplementary Figure S1) were considered as physical WCT location. This enabled the analysis of the temporal evolution of WCT on the body surface. To characterize the overall spatial distribution of WCT during the P-wave, for each point *x*, the percentage of time *p* during which the absolute difference of WCT and BSPM was smaller than the threshold *τ*, was computed by

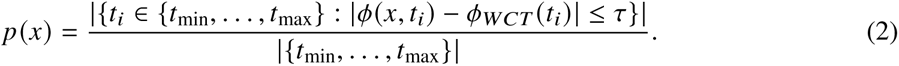

The resulting spatial distributions of *p* were visually inspected and manually classified. Additionally, WCT was compared to the BSPM signals at the center of the Einthoven triangle *ϕ*_ET_(*t*) and the center between the V1 and V2 electrode positions *ϕ*_V1___V2_(*t*). Agreement between these signals and WCT was further assessed using Equation 2 for *ϕ*_ET_ and *ϕ*_V1___V2_.

**Table 2.** Parameters for the electrophysiological simulations. Conduction velocities and anisotropy ratios for seven distinct regions in the atria were chosen as in previous studies [1].

| Atrial region | Conduction velocity | Anisotropy ratio |
| --- | --- | --- |
| Interatrial connections | 1093 mm/s | 3.36 |
| Right atrium | 739 mm/s | 2.11 |
| Left atrium | 946 mm/s | 2.11 |
| Valve rings | 445 mm/s | 2.11 |
| Pectinate muscles | 578 mm/s | 3.78 |
| Crista terminalis | 607 mm/s | 3.00 |
| Inferior isthmus | 722 mm/s | 1.00 |

### A.2 Clinical Data on WCT Location

For the clinical analysis, 10 s segments of previously acquired BSPMs from 10 normal subjects [13] were analyzed (Table 1). All clinically recorded BSPMs were referenced to the RL and noisy signals were semi-automatically identified and removed [13]. A template of the P-wave was generated [14] and bandpass (0.05-40 Hz) filtered. The P-wave was annotated manually based on the 12-lead ECG template, and the BSPM signals were corrected for isoline offset. Each torso model derived from the CT scan was remeshed to an average mesh resolution of 15 mm similar to the in silico models using *InstantMeshes* [15] and the iso2mesh Toolbox [16] and smoothed using 200 iterations [17]. Laplacian interpolation was applied to interpolate the BSPM signals to the CT scan of the torso surface [12]. For the clinical study, limb electrodes were chosen based on the nearest electrode that was available for each position on the torso. As clinical signal amplitudes were smaller compared simulated data, an amplitude-adjusted threshold was tested. As this yielded, similar, though less distinct, spatial patterns (Supplementary Figure S1), the same fixed threshold was used for both setups to ensure consistency.

**Figure S1.**
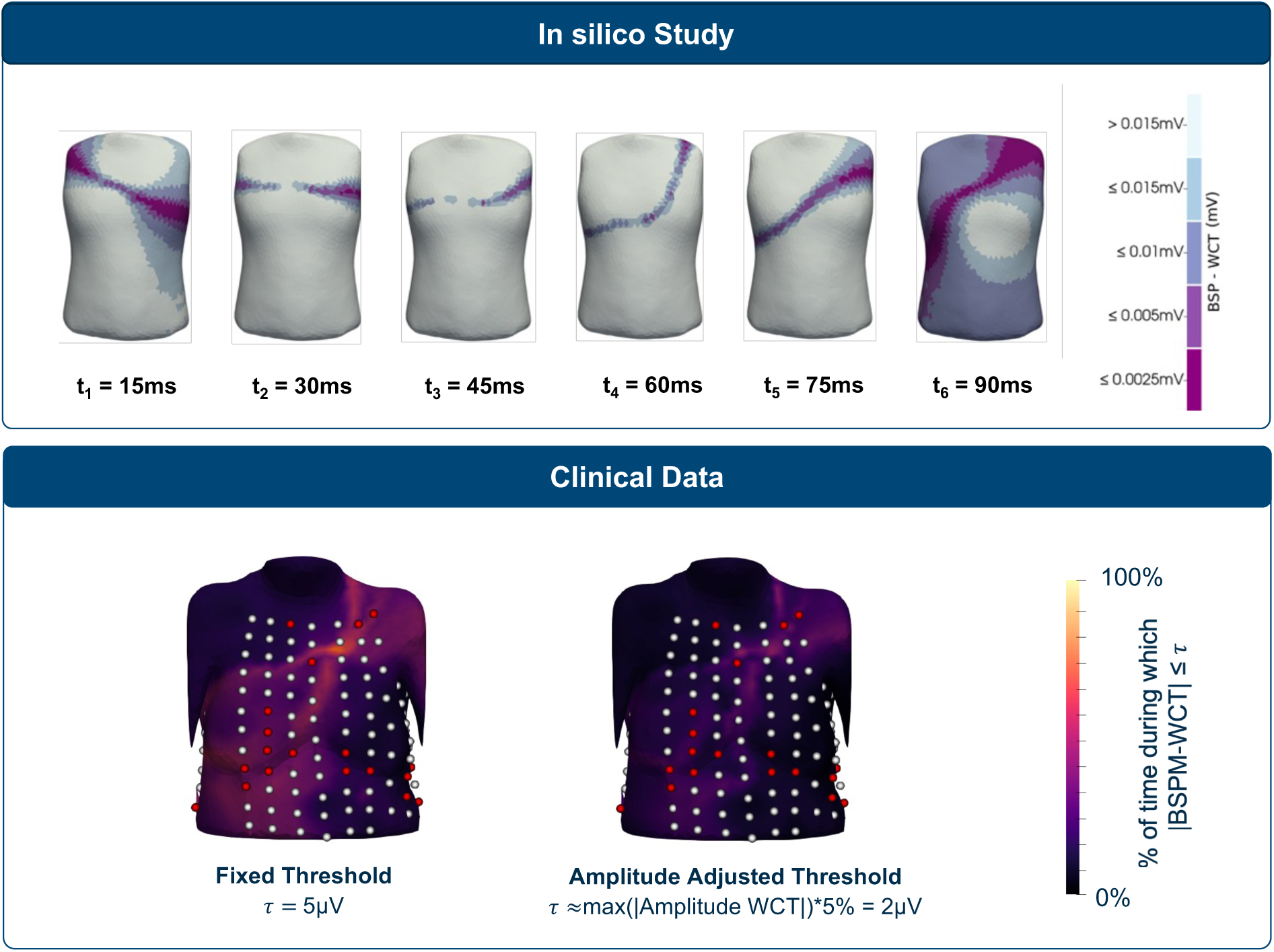
**Top** Evaluation of different thresholds for Equation 2 for the mean atrial model and mean torso model. The larger the threshold, the larger the area in which Equation 2 is fulfilled. For this study, a fixed threshold of 5 *μV* was chosen. **Bottom** Threshold for Equation 2 in the clinical cohort. Using the same fixed threshold as in the in silico setup, larger percentages were observed due to smaller signal amplitudes (left). An amplitude adjusted threshold of 5% of maximum absolute WCT amplitude lead to smoothed patterns, slightly smaller percentages for most models and overall similar spatial patterns. Electrodes of the clinical BSPM are visualized as spheres and electrodes with noisy signals that were interpolated in red. WCT: Wilson’s central terminal, BSPM: Body surface potential map.

## B Supplementary Figures

**Figure S2.**
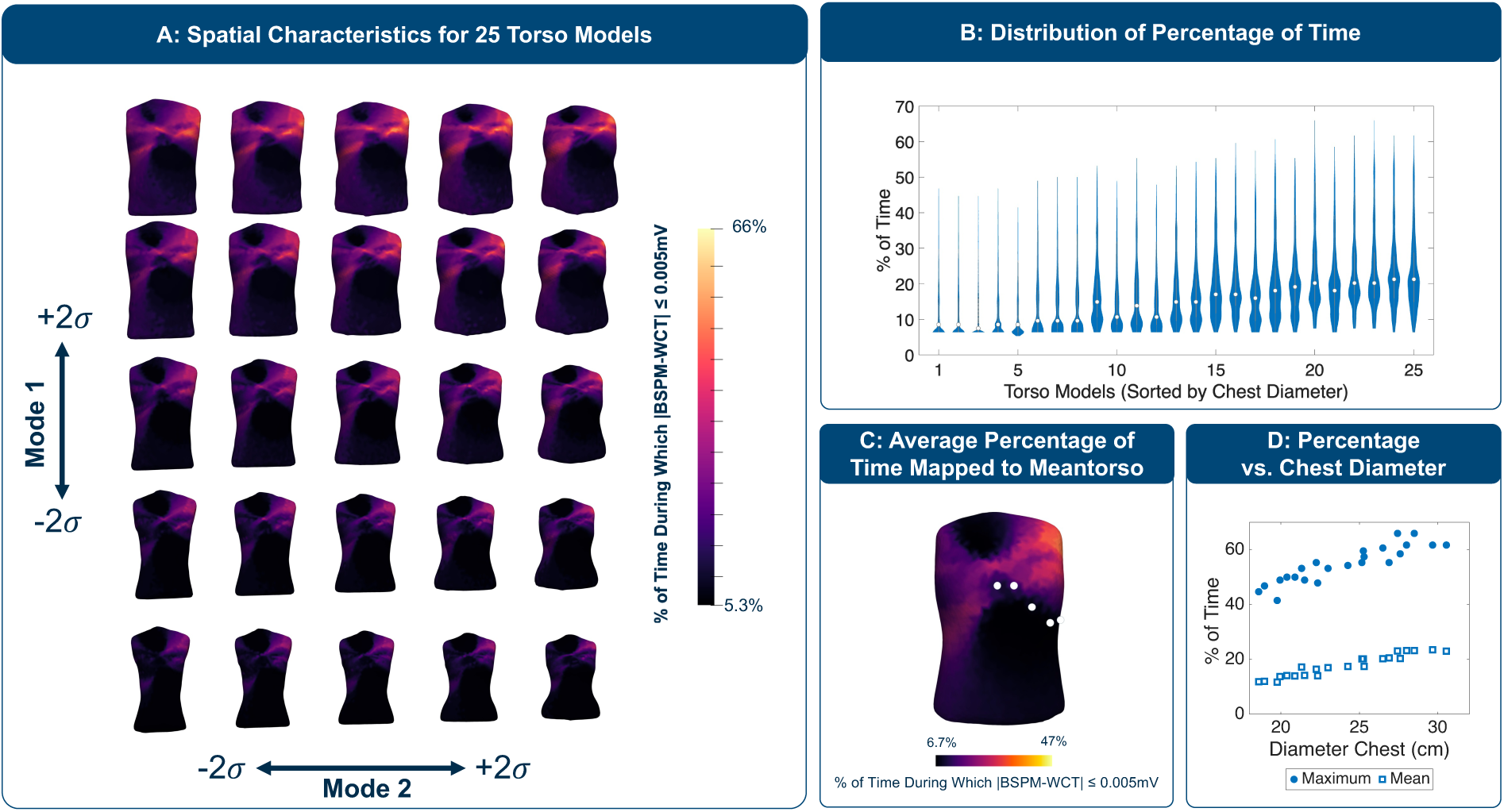
Differences in WCT location for different models. **A:** Spatial patterns of WCT on the body surface for different torso models and the mean atrial model. Torso models are ordered according to the coefficients of the two first eigenmodes that were varied systematically. The mean torso is located in the center. Abdominal diameter decreases from top to bottom (1st eigenmode), chest diameter increases from left to right in each row (2nd eigenmode). Volume and surface area of the torso models decrease from top to bottom and from left to right. **B:** Violin plots showing the distribution of percentage of time during which the difference of BSPM and WCT is ≤ 0.005 mV, calculated across all points on the body surface for each torso model. Torso models sorted by increasing chest diameters. **C:** Spatial distribution of the mean percentage of time across all 25 torso models after mapping to the mean torso model. **D:** Maximum and mean percentages of time for different torso models and mean atrial model, sorted by chest diameter. WCT: Wilson’s central terminal, BSPM: Body surface potential map.

**Figure S3.**
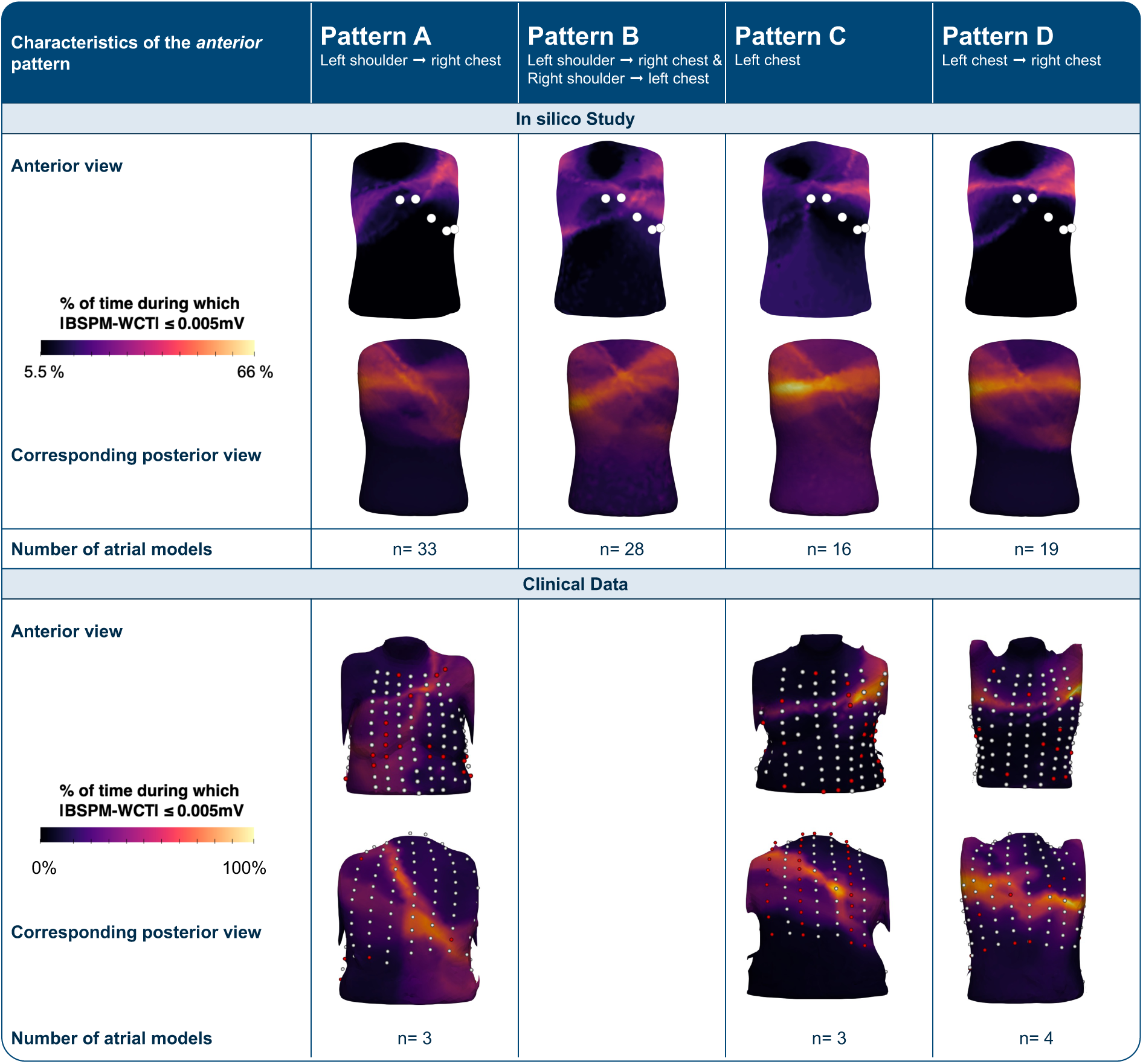
Spatial patterns of WCT on the body surface, anterior and posterior views. The anterior patterns were manually classified into four different categories according to the spatial extension and location of WCT on the body surface. Posterior patterns are shown for the corresponding example geometry that was chosen to visualize the anterior pattern. Small deviations between WCT and BSPM signals were found posteriorly due to overall lower posterior amplitudes. In the clinical cohort, no example for pattern B was found. Spheres represent electrode positions of the BSPM, and electrodes that were to noisy and thus not used in the analysis are visualized in red. 3 out 99 models could not be assigned to one of the defined patterns but had large percentages in the anterior abdominal region and posteriorly in the medial superior chest. WCT: Wilson’s central terminal, BSPM: Body surface potential map.

**Figure S4.**
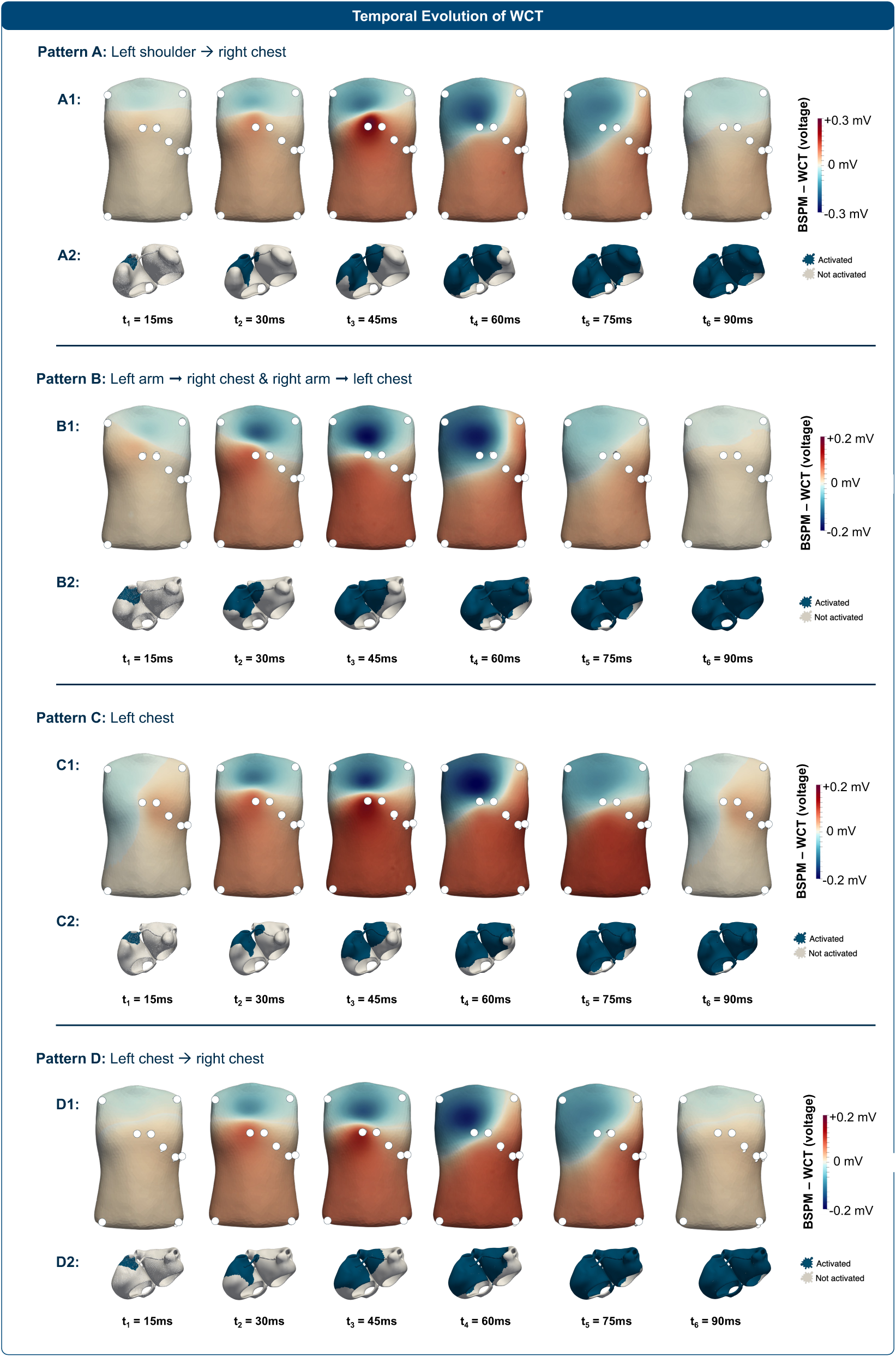
**A1, B1, C1, D1:** Temporal evolution of the WCT location on the body surface for the four spatial patterns of WCT. Electrode positions for the extremities and the leads V1-V5 are indicated by white spheres. White areas on the body surface indicate regions where the absolute difference between WCT and the BSPM was ≤ 5 *μV* and can thus be considered as WCT location. **A2, B2, C2, D2:** Depolarized regions of the atria at each time point are shown in blue. WCT: Wilson’s central terminal, BSPM: Body surface potential map.

## References

[1] Frank N Wilson et al. “Electrocardiograms That Represent the Potential Variations of a Single Electrode”. In: American Heart Journal 9.4 (1934). doi: 10.1016/S0002-8703(34)90093-4.

[2] Mario J. Mc Loughlin and Pedro Brugada. “Electrodes and Leads in Electrocardiography: A Conceptual Review”. In: Current Problems in Cardiology 50.10 (Oct. 2025), p. 103150. issn: 01462806. doi: 10.1016/j.cpcardiol.2025.103150.

[3] Hossein Moeinzadeh et al. “Unipolar Cardiac Leads Between History and Science”. In: Biomedical Signal Processing. Ed. by Ganesh Naik. Singapore: Springer Singapore, 2020, pp. 203–224. doi: 10.1007/978-981-13-9097-5_10.

[4] H.C. Burger. “The Zero of Potential: A Persistent Error”. In: American Heart Journal 49.4 (Apr. 1955), pp. 581–586. issn: 00028703. doi: 10.1016/0002-8703(55)90076-4.

[5] G. E. Dower, J. A. Osborne, and A. D. Moore. “Measurement of the error in Wilson’s central terminal: an accurate definition of unipolar leads”. In: Heart 21.3 (July 1959), pp. 352–360. issn: 1355-6037. doi: 10.1136/hrt.21.3.352. (Visited on 10/02/2025).

[6] Frank N. Wilson, A. Garrard Macleod, and Paul S. Barker. “The Potential Variations Produced by the Heart Beat at the Apices of Einthoven’s Triangle”. In: American Heart Journal 7.2 (Dec. 1931), pp. 207–211. issn: 00028703. doi: 10.1016/S0002-8703(31)90411-0.

[7] Norio Miyamoto et al. “The Absolute Voltage and the Lead Vector of Wilson’s Central Terminal.” In: Japanese Heart Journal 37.2 (1996), pp. 203–214. issn: 0021-4868, 1348-673X. doi: 10.1536/ ihj.37.203.

[8] Yoshiwo Okamoto and Saburo Mashima. “The Zero Potential and Wilson’s Central Terminal in Electrocardiography”. In: Bioelectrochemistry and Bioenergetics 47.2 (Dec. 1998), pp. 291–295. issn: 03024598. doi: 10.1016/S0302-4598(98)00201-3.

[9] Hossein Moeinzadeh et al. “Minimization of the Wilson’s Central Terminal Voltage Potential via a Genetic Algorithm”. In: BMC Research Notes 11.1 (Dec. 2018), p. 915. issn: 1756-0500. doi: 10.1186/s13104-018-4017-y.

[10] Frank N. Wilson et al. “Recommendations for Standardization of Electrocardiographic and Vectorcardiographic Leads”. In: Circulation 10.4 (Oct. 1954), pp. 564–573. issn: 0009-7322, 1524-4539. doi: 10.1161/01.CIR.10.4.564.

[11] Gaetano D. Gargiulo. “True Unipolar ECG Machine for Wilson Central Terminal Measurements”. In: BioMed Research International 2015 (2015), pp. 1–7. issn: 2314-6133, 2314-6141. doi: 10.1155/2015/586397.

[12] Ljuba Bacharova et al. “Where Is the Central Terminal Located?” In: Journal of Electrocardiology 38.2 (Apr. 2005), pp. 119–127. issn: 00220736. doi: 10.1016/j.jelectrocard.2005.01.002.

[13] Jaakko Malmivuo and Robert Plonsey. Bioelectromagnetism Principles and Applications of Bioelectric and Biomagnetic Fields. Oxford University Press, Oct. 1995. doi: 10.1093/acprof:oso/9780195058239.001.0001.

[14] Hossein Moeinzadeh et al. “WCTECGdb: A 12-Lead Electrocardiography Dataset Recorded Simultaneously with Raw Exploring Electrodes’ Potential Directly Referred to the Right Leg”. In: Sensors 20.11 (June 2020), p. 3275. issn: 1424-8220. doi: 10.3390/s20113275.

[15] Gaetano D. Gargiulo et al. “On the Einthoven Triangle: A Critical Analysis of the Single Rotating Dipole Hypothesis”. In: Sensors 18.7 (July 2018), p. 2353. issn: 1424-8220. doi: 10.3390/s18072353.

[16] Pyotr G. Platonov. “Atrial conduction and atrial fibrillation: What can we learn from surface ECG?” In: Cardiology Journal 15.5 (2008), pp. 402–407. issn: 1898-018X.

[17] Mahesh Kumar Batra et al. “Assessment of Electrocardiographic Criteria of Left Atrial Enlargement”. In: Asian Cardiovascular and Thoracic Annals 26.4 (May 2018), pp. 273–276. issn: 0218-4923, 1816-5370. doi: 10.1177/0218492318768131. (Visited on 12/02/2025).

[18] Junrong Jiang et al. “Detection of Left Atrial Enlargement Using a Convolutional Neural Network-Enabled Electrocardiogram”. In: Frontiers in Cardiovascular Medicine 7 (Dec. 2020), p. 609976. issn: 2297-055X. doi: 10.3389/fcvm.2020.609976.

[19] Mark S Hazen, Thomas H Marwick, and Donald A Underwood. “Diagnostic accuracy of the resting electrocardiogram in detection and estimation of left atrial enlargement: an echocardiographic correlation in 551 patients”. In: American heart journal 122.3 (1991), pp. 823–828.

[20] Emma Julia Petronella Nilsson et al. “ECG and CT for the Detection of Left Atrial Enlargement in Hypertensive Individuals—a Population-Based Study”. In: Hypertension Research 45.8 (Aug. 2022), pp. 1382–1391. issn: 0916-9636, 1348-4214. doi: 10.1038/s41440-022-00918-z.

[21] Robin Andlauer et al. “Influence of left atrial size on P-wave morphology: differential effects of dilation and hypertrophy”. In: Europace 20.S3 (Nov. 2018), pp. iii36–iii44. doi: 10.1093/europace/euy231.

[22] Pyotr G. Platonov. “P-Wave Morphology: Underlying Mechanisms and Clinical Implications”. In: Annals of Noninvasive Electrocardiology 17.3 (2012), pp. 161–169. doi: 10.1111/j.1542-474X.2012.00534.x.

[23] Job Stoks et al. “Variant Patterns of Electrical Activation and Recovery in Normal Human Hearts Revealed by Noninvasive Electrocardiographic Imaging”. In: Europace 26.7 (July 2024). issn: 1099-5129, 1532-2092. doi: 10.1093/europace/euae172.

[24] Claudia Nagel et al. “A Bi-Atrial Statistical Shape Model for Large-Scale in Silico Studies of Human Atria: Model Development and Application to ECG Simulations”. In: Medical Image Analysis 74 (Dec. 2021), p. 102210. issn: 13618415. doi: 10.1016/j.media.2021.102210.

[25] Leonid Pishchulin et al. “Building Statistical Shape Spaces for 3D Human Modeling”. In: Pattern Recognition 67 (July 2017), pp. 276–286. issn: 00313203. doi: 10.1016/j.patcog.2017.02.018.

[26] Claudia Nagel et al. “Non-Invasive and Quantitative Estimation of Left Atrial Fibrosis Based on P Waves of the 12-Lead ECG—A Large-Scale Computational Study Covering Anatomical Variability”. In: Journal of Clinical Medicine 10.8 (Apr. 2021), p. 1797. issn: 2077-0383. doi: 10.3390/jcm10081797.

[27] Claudia Nagel et al. “Comparison of Propagation Models and Forward Calculation Methods on Cellular, Tissue and Organ Scale Atrial Electrophysiology”. In: IEEE Transactions on Biomedical Engineering 70.2 (Feb. 2023), pp. 511–522. issn: 0018-9294, 1558-2531. doi: 10.1109/TBME.2022.3196144.

[28] Silvia Becker et al. “Deep Learning for Amplified P-Wave Duration Annotation”. In: Proceedings of the 52nd Computing in Cardiology Conference (CinC 2025). 2025, p. 1.

[29] Ioannis Vogiatzis et al. “The Importance of the 15-lead Versus 12-lead ECG Recordings in the Diagnosis and Treatment of Right Ventricle and Left Ventricle Posterior and Lateral Wall Acute Myocardial Infarctions”. In: Acta Informatica Medica 27.1 (Jan. 2019), p. 35. doi: 10.5455/aim.2019.27.35-39.

[30] Giorgio Luongo et al. “Automatic ECG-based Discrimination of 20 Atrial Flutter Mechanisms: Influence of Atrial and Torso Geometries”. In: 2020 Computing in Cardiology. 9344051. IEEE, Jan. 2020, pp. 1–4. doi: 10.22489/CinC.2020.066.

[31] John Silberbauer. “Wilson’s Central Terminal, the Keystone to Electrogram Recording – What, Where and Why?” In: (2013). (Visited on 08/19/2025).

[32] Claudia Nagel et al. [dataset] A Bi-atrial Statistical Shape Model and 100 Volumetric Anatomical Models of the Atria. June 2021. doi: 10.5281/ZENODO.5571925. url: https://doi.org/10.5281/zenodo.4309957.

## References

[1] Claudia Nagel et al. “A Bi-Atrial Statistical Shape Model for Large-Scale in Silico Studies of Human Atria: Model Development and Application to ECG Simulations”. In: Medical Image Analysis 74 (Dec. 2021), p. 102210. issn: 13618415. doi: 10.1016/j.media.2021.102210.

[2] Claudia Nagel et al. [dataset] A Bi-atrial Statistical Shape Model and 100 Volumetric Anatomical Models of the Atria. June 2021. doi: 10.5281/ZENODO.5571925. url: https://doi.org/10.5281/zenodo.4309957.

[3] Luca Azzolin et al. “AugmentA: Patient-specific Augmented Atrial Model Generation Tool”. In: Computerized Medical Imaging and Graphics 108 (Sept. 2023), p. 102265. issn: 08956111. doi: 10.1016/j.compmedimag.2023.102265.

[4] Leonid Pishchulin et al. “Building Statistical Shape Spaces for 3D Human Modeling”. In: Pattern Recognition 67 (July 2017), pp. 276–286. issn: 00313203. doi: 10.1016/j.patcog.2017.02.018.

[5] Claudia Nagel et al. “Non-Invasive and Quantitative Estimation of Left Atrial Fibrosis Based on P Waves of the 12-Lead ECG—A Large-Scale Computational Study Covering Anatomical Variability”. In: Journal of Clinical Medicine 10.8 (Apr. 2021), p. 1797. issn: 2077-0383. doi: 10.3390/jcm10081797.

[6] Claudia Nagel et al. “Comparison of Propagation Models and Forward Calculation Methods on Cellular, Tissue and Organ Scale Atrial Electrophysiology”. In: IEEE Transactions on Biomedical Engineering 70.2 (Feb. 2023), pp. 511–522. issn: 0018-9294, 1558-2531. doi: 10.1109/TBME.2022.3196144.

[7] A. Loewe et al. “Influence of the earliest right atrial activation site and its proximity to interatrial connections on P-wave morphology”. In: Europace 18.suppl 4 (Jan. 2016), pp. iv35–iv43.

[8] J.A. Sethian. Level Set Methods and Fast Marching Methods. Aug. 1999. isbn: 978-0-521-64557-7.

[9] Cristian Barrios Espinosa et al. “A cyclical fast iterative method for simulating reentries in cardiac electrophysiology using an eikonal-based model”. In: Engineering with Computers (Jan. 2025). doi: 10.1007/s00366-024-02094-9.

[10] Marc Courtemanche, Rafael J. Ramirez, and Stanley Nattel. “Ionic Mechanisms Underlying Human Atrial Action Potential Properties: Insights from a Mathematical Model”. In: American Journal of Physiology-Heart and Circulatory Physiology 275.1 (July 1998), H301–H321. issn: 0363-6135, 1522-1539. doi: 10.1152/ajpheart.1998.275.1.H301.

[11] David U J Keller et al. “Ranking the Influence of Tissue Conductivities on Forward-Calculated ECGs”. In: IEEE Transactions on Biomedical Engineering 57.7 (July 2010), pp. 1568–1576. issn: 0018-9294. doi: 10.1109/TBME.2010.2046485.

[12] Steffen Schuler et al. “Spatial Downsampling of Surface Sources in the Forward Problem of Electrocardiography”. In: Functional Imaging and Modeling of the Heart. Ed. by Yves Coudière et al. Vol. 11504. Cham: Springer International Publishing, 2019, pp. 29–36. doi: 10.1007/978-3-030-21949-9_4.

[13] Job Stoks et al. “Variant Patterns of Electrical Activation and Recovery in Normal Human Hearts Revealed by Noninvasive Electrocardiographic Imaging”. In: Europace 26.7 (July 2024). issn: 1099-5129, 1532-2092. doi: 10.1093/europace/euae172.

[14] Silvia Becker et al. “Deep Learning for Amplified P-Wave Duration Annotation”. In: Proceedings of the 52nd Computing in Cardiology Conference (CinC 2025). 2025, p. 1.

[15] Wenzel Jakob et al. “Instant Field-Aligned Meshes”. In: ACM Transactions on Graphics 34.6 (Nov. 2015), pp. 1–15. issn: 0730-0301, 1557-7368. doi: 10.1145/2816795.2818078.

[16] Anh Phong Tran, Shijie Yan, and Qianqian Fang. “Improving Model-Based Functional near-Infrared Spectroscopy Analysis Using Mesh-Based Anatomical and Light-Transport Models”. In: Neurophotonics 7.01 (Feb. 2020), p. 1. issn: 2329-423X. doi: 10.1117/1.NPh.7.1.015008.

[17] James Ahrens, Berk Geveci, and Charles Law. “Paraview: An End-User Tool for Large Data Visualization”. In: The visualization handbook 717.8 (2005).

